# Analysis of MRI-derived spleen iron in the UK Biobank identifies genetic variation linked to iron homeostasis and erythrocyte morphology

**DOI:** 10.1101/2021.11.16.21266431

**Authors:** Elena P. Sorokin, Nicolas Basty, Brandon Whitcher, Yi Liu, Jimmy D. Bell, Robert L. Cohen, Madeleine Cule, E. Louise Thomas

**Author notes:** These authors contributed equally (Elena P. Sorokin, Nicolas Basty, Madeleine Cule, E. Louise Thomas).

## Abstract

The spleen plays a key role in iron homeostasis. It is the largest filter of the blood and performs iron reuptake from old or damaged erythrocytes. Despite this role, spleen iron concentration has not been measured in a large, population-based cohort. In this study, we quantify spleen iron in 41,764 participants of the UK Biobank using magnetic resonance imaging, and provide the first reference range for spleen iron in an unselected population. Through genome-wide association study, we identify associations between spleen iron and regulatory variation at two hereditary spherocytosis genes, *ANK1* and *SPTA1*. Spherocytosis-causing coding mutations in these genes are associated with lower reticulocyte volume and increased reticulocyte percentage, while these novel common alleles are associated with increased expression of *ANK1* and *SPTA1* in blood and with larger reticulocyte volume and reduced reticulocyte percentage. As genetic modifiers, these common alleles may explain mild spherocytosis phenotypes that have been observed clinically. Our genetic study also identifies a signal which co-localizes with a splicing quantitative trait locus for *MS4A7*, and we show this gene is abundantly expressed in the spleen and in macrophages. The combination of deep learning and efficient image processing enables non-invasive measurement of spleen iron and, in turn, characterization of genetic factors related to iron recycling and erythrocyte morphology.

## Introduction

In normal human physiology, iron is recycled much faster than new dietary iron is absorbed, and this iron economy is regulated by the spleen.^1–3^ The spleen plays a critical role in removing senescent erythrocytes from the blood, and does so via a population of splenic macrophages as well as by the action of the protein ferroportin, which transports iron back to the plasma.^4^ Given the large iron flux through the normal spleen due to erythrocyte recycling, measurement of spleen iron has the potential to reflect activity of iron salvage pathways.

Spleen iron also may reflect erythrocyte biology and dysfunction. Hereditary spherocytosis (HS) is a relatively common erythrocyte membrane disorder, occurring at a prevalence of 1:1000-2500 in European populations.^5^ Patients have defects in membrane and cytoskeletal genes that contribute to erythrocyte membrane integrity and deformability, including *SPTA1 (*encoding the filamentous protein alpha-spectrin) and *ANK1 (*encoding a protein that tethers spectrin filaments to erythrocyte membranes).^*6,7*^ The protein products of these genes interact in the formation of the mature erythrocyte cytoskeleton, a process accompanying cellular remodeling during reticulocytosis.^8^ In HS, the resulting erythrocytes are spherically shaped and lose deformability, and are ultimately trapped and ingested by macrophages within the red pulp of the spleen, resulting in an enlarged spleen and anemia despite brisk reticulocytosis. When measured directly with radiolabeled cells, erythrocyte turnover is dramatically accelerated in patients with severe HS.^9^ While laboratory measures such as sphered cell and reticulocyte volume can diagnose HS^10^, less is understood about the phenotypic and genetic heterogeneity of the disease.^11–13^

Although spleen iron has been investigated in specific disease groups^14–20^, few studies were performed in unascertained cohorts^21^ partly due to limitations in detecting and quantifying low levels of tissue iron in non-overloaded populations.^22,23^ The UK Biobank (UKBB) is a prospective study of half a million adults in the UK,^24^ with genetic and phenotypic data including magnetic resonance imaging (MRI).^25^ In this study, we applied computer vision techniques to quantify spleen iron non-invasively and at scale, by repurposing the dedicated liver MRI acquisition. Spleen iron is only moderately correlated with other measures of iron stores in the body, making it a novel trait. Through genome-wide association study (GWAS), we characterized associations between spleen iron and genes involved in the human iron economy including common regulatory variation in the ferroportin gene, *SLC40A1*. Our GWAS identified a signal which co-localizes with a splicing quantitative trait locus for *MS4A7*, which we found to be abundantly expressed in the spleen and in macrophages. Regulatory variation in the genes encoding alpha-spectrin (*SPTA1*) and ankyrin (*ANK1*) were also linked to spleen iron, increased mRNA expression of these genes, and furthermore exhibited effects on reticulocyte and erythrocyte parameters opposite to the effects observed in HS patients. Non-invasive imaging coupled with genetic analysis enabled us to develop a novel trait and illuminated new aspects of iron homeostasis and erythrocyte biology.

## Results

### Characterization of spleen iron in a large, population-based cohort

We quantified spleen iron concentration (spleen iron hereafter) in 41,764 UK Biobank (UKBB) participants with both the 3D neck-to-knee and the quantitative liver single-slice MRI sequences available. We opportunistically measured spleen iron concentration by segmenting the spleen from the neck-to-knee image^26^ (**Figure 1A**) and subsequently extracting a 2D mask where the spleen volume intersects with the quantitative 2D liver slice (**Figure 1B**).^45^ The average spleen iron was 0.92±0.32 mg/g, significantly lower than liver iron of 1.24±0.29 mg/g (**Table 1, Figure 1C**; paired t-test p < 2.2e-16). While there is no accepted normal range of spleen iron, 1.98 mg/g has been suggested as an upper cut-off, with 2.74 mg/g reported to be pathological.^27^ Using 1.98 mg/g as the threshold, 1.04% (n=435) of this cohort had elevated spleen iron, while 0.32% (n=137) had spleen iron above the 2.74 mg/g threshold. 95% of the population fell into the range of 0.53 to 1.67 mg/g, and we propose this as a possible reference range in an unselected population.

**Figure 1:**
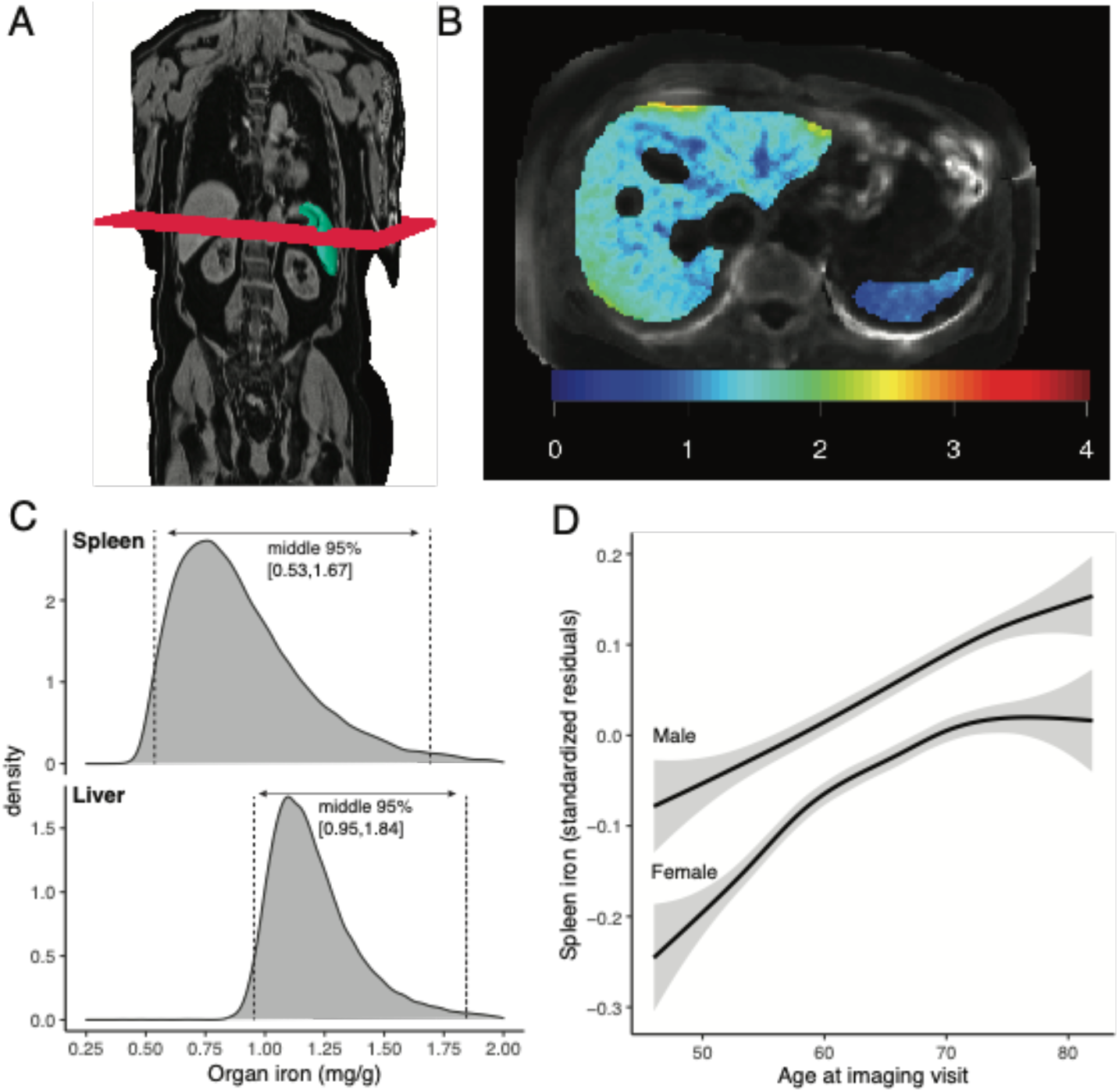
High-throughput spleen iron quantification. **A)** Example neck-to-knee abdominal MRI acquisition, with the liver slice acquistion shown as a red plane, and example deep learning-derived spleen segmentation shown in green. **B)** Axial view of iron concentration (mg/g) obtained through deep learning and image processing, with liver shown at left and the spleen at right. **C)** Distribution of spleen iron (mg/g), as compared to liver iron, in the UK Biobank (n=41,764). **D)** Spleen iron increases with age for both males and females. Standardized residuals are shown, adjusted for study center, scan date and scan time.

**Table 1:** Summary of the UKBB cohort at recruitment, image acquisition, and quantification of liver iron and spleen iron. Spleen iron measurements (mg/g) are provided as mean (standard deviation) and range. For other quantitative values, the mean (standard deviation) is given. For binary values, mean [95% confidence interval] is given.

|  | UK Biobank cohort (at time of baseline visit) | Imaging cohort (at time of imaging visit) | Identified in combined liver slices |  |
| --- | --- | --- | --- | --- |
|  |  |  | Liver | Spleen |
| n | 502,520 | 44,265 | 44,265 | 41,764 |
| % Female | 54.4 [54.2-54.5] | 51.8 [51.2-52.1] | 51.8 [51.3-52.3] | 52.1 [51.6-52.6] |
| Age | 56.5 (8.1) | 64.2 (7.73) | 63.7 (7.56) | 64.1 (7.71) |
| BMI (kg/m<sup>2</sup>) | 27.4 (4.8) | 26.5 (4.37) | 26.5 (4.39) | 26.4 (4.32) |
| Height (cm) | 168 (9.28) | 169 (9.27) | 169 (9.29) | 169 (9.26) |
| % Caucasian | 81.4 [81.3-81.5] | 84.8 [84.4-85.1] | 85.2 [84.4-85.5] | 84.7 [84.4-85.0] |
| Iron concentration (mg/g) |  |  | 1.24 (0.29) [0.203 - 6.96] | 0.915 (0.318) [0.172 - 6.68] |

Spleen iron differed by age and sex. Men had higher spleen iron than women (men: 0.96±0.34 mg/g, women: 0.87±0.29 mg/g) (p=6.3e-219; **Supplementary Table 2**). Increasing spleen iron was associated with age (0.0044 mg/g/year or 0.012 SD/year) (**Figure 1D**). In women, menopause was associated with 0.12 mg/g higher spleen iron [95% CI 0.08-0.16].

In a phenome-wide association study with over 3,200 quantitative traits and disease outcomes, spleen iron was correlated with erythrocyte parameters: reticulocyte percentage (beta=0.091; p=3.7e-66), reticulocyte count (beta=0.087; p=6.6e-64), and high light scatter reticulocyte count (beta=0.089; p=5.8e-64). Spleen iron was also associated with lifestyle factors, including consumption of lamb (beta=0.150; p=2.1e-44) and beef (beta=0.143; p=1.3e-43), and negatively associated with alcohol consumption (beta=-0.087; p=1.7e-18). Spleen iron correlated with brain iron content, specifically with T2* (inversely proportional to iron) in the caudate (beta=-0.063; p=4.3e-16) and putamen (beta=-0.061, p=4.1e-15).^28^ Spleen iron was associated with myeloid leukemia (beta=0.386; p=9.6e-10), chronic dermatitis (beta=0.328; p=4.1e-07), hypokalemia (beta =0.286, p=5.8e-6), and glaucoma (beta=0.186, p=1.0e-6) (**Figure 2, Supplementary Tables 3-4**). Spleen iron was negatively correlated with iron-deficiency anemia, but this did not achieve Bonferroni significance (beta=-0.134, p=0.002), though this diagnosis may not have been fully captured by medical billing codes (n=580 cases, n=35,316 controls).

**Figure 2:**
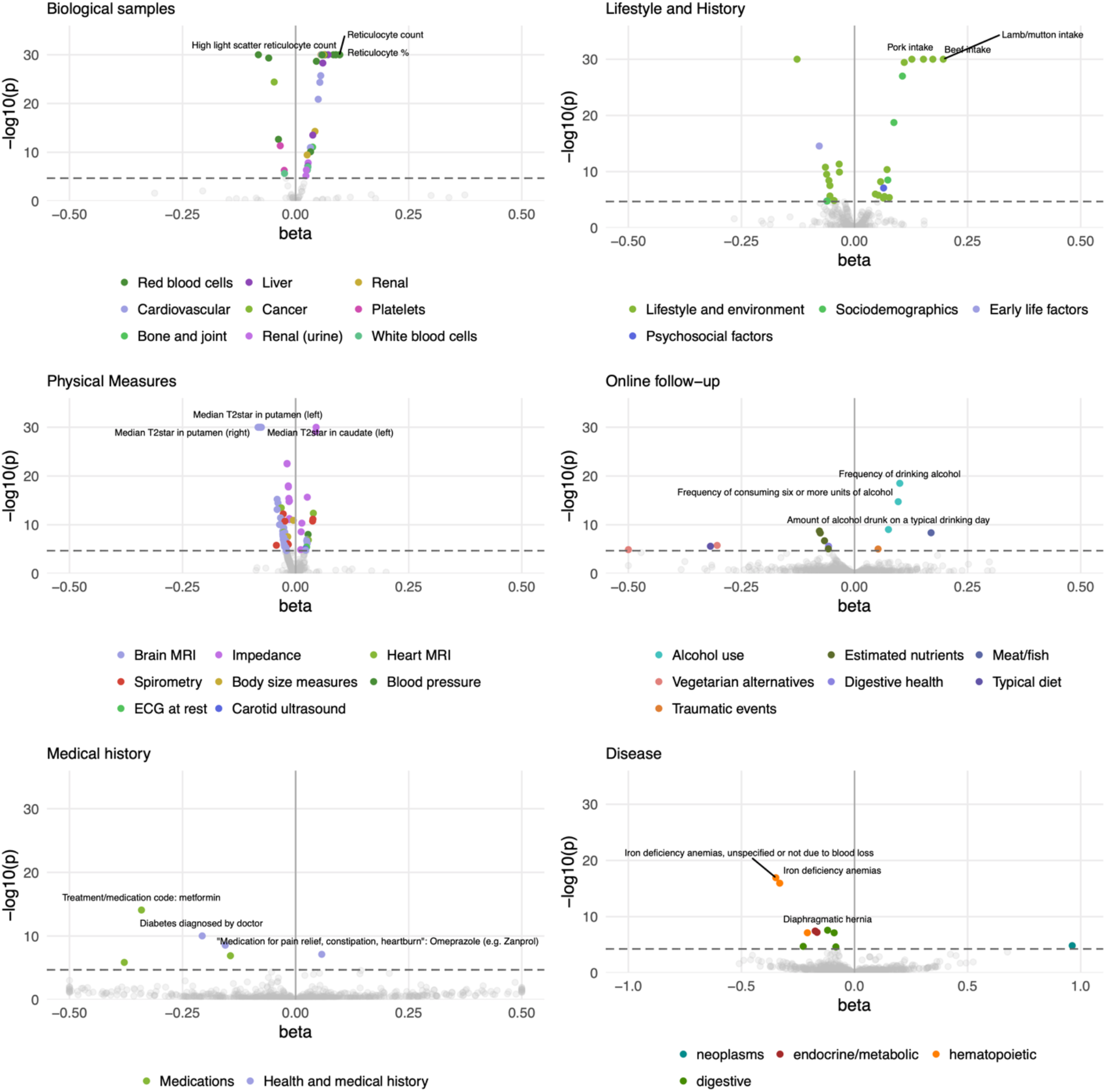
Phenome-wide association between spleen iron and complex traits in the UK Biobank. The horizontal line shows a significant association after Bonferroni correction. The top three associations in each category are annotated. ML: Myeloid leukaemia. HLS reticulocyte: High light scatter reticulocyte.

### Genome-wide association study of spleen iron identifies DNA polymorphisms linked to global iron homeostasis

In a common variant genome-wide association study (GWAS) of spleen iron, seven loci reached genome-wide significance (p < 5e-8; **Figure 3, Table 2**). Conditional analysis yielded no secondary signals. We estimated the narrow-sense heritability of spleen iron to be 16.7% (s.e. 1.64%). Spleen iron was moderately genetically correlated with ferritin (r_g_=0.56) and mean corpuscular hemoglobin concentration (r_g_=0.42), but the genetic correlations with other iron measures, including liver iron, were not significant (**Supplementary Tables 5-6)**.

**Figure 3.**
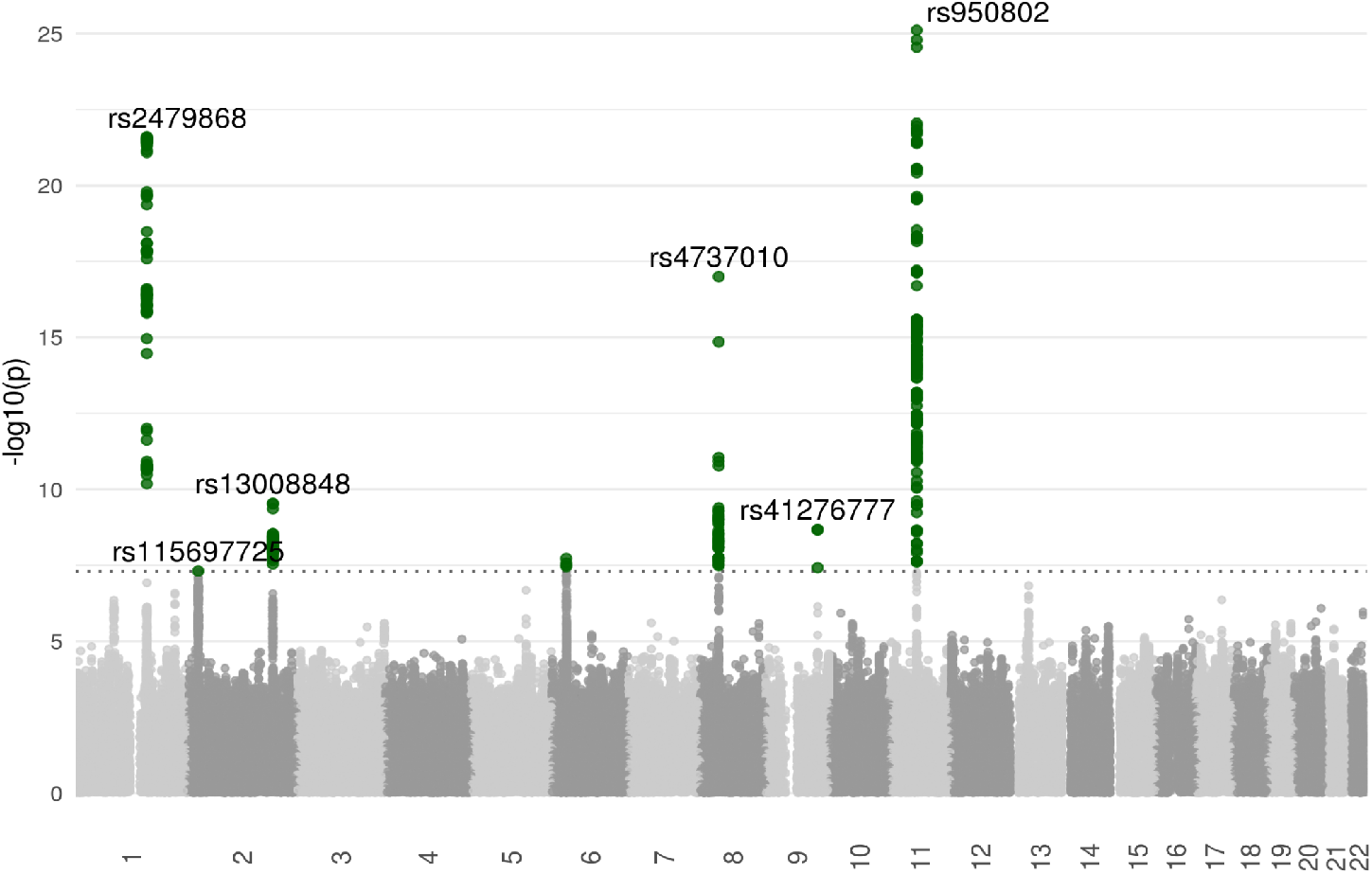
Genome-wide association study of spleen iron. Genome-wide significant signals are annotated with the variant at each locus. Dotted horizontal line marks genome-wide significance (p=5e-8).

**Table 2:** Fine-mapped lead SNPs from genome-wide association study of spleen iron. Genome-wide significant associations (p<5e-8) are shown by locus and lead SNP after fine-mapping. A seventh association at the major histocompatibility locus (MHC) could not be fine-mapped.

| Lead SNP | Locus | Lead SNP consequence | Effect Allele | Other Allele | Beta | p | Minor Allele Frequency |
| --- | --- | --- | --- | --- | --- | --- | --- |
| rs950802 | *MS4A7/MS4A14* | Synonymous variant | A | G | 0.086 | 7.7E-26 | 0.307 |
| rs2479868 | *SPTA1* | Intron variant | T | C | -0.083 | 2.5E-22 | 0.266 |
| rs4737010 | *ANK1* | Intron variant | A | G | -0.077 | 1.0E-17 | 0.228 |
| rs13008848 | *SLC40A1* | 5 prime UTR variant | G | C | -0.057 | 2.9E-10 | 0.226 |
| rs41276777 | *PRPF4* | 5 prime UTR variant | A | G | 0.175 | 2.1E-09 | 0.019 |
| rs115697725 | *KLHL29* | Intron variant | G | C | 0.059 | 4.9E-08 | 0.146 |

We observed a signal on chromosome 2 at *SLC40A1*, which encodes the iron transporter ferroportin (lead variant: rs13008848[G], beta=-0.057, p=2.9e-10, **Figure 4A**). To test whether the spleen signal was shared across other body iron traits, we re-analyzed previous genetic studies of serum ferritin, serum iron, and liver iron^26,29^, and observed evidence of co-localization at *SLC40A1*(posterior probability≥0.99) (**Supplementary Table 7**). Since the lead variant lies upstream of the *SLC40A1* open reading frame, we tested for a shared effect between spleen iron and *SLC40A1* expression. Across 56 human tissues of the GTEx Consortium,^30^ we observed evidence for regional co-localization with a quantitative trait locus for *SLC40A1* mRNA in many tissues, including whole blood (posterior probability≥0.99) (**Figure 4D**; **Supplementary Table 8**). As expected, spleen iron was associated with *SLC40A1*, and furthermore, we showed that this locus likely influences tissue iron levels through *SLC40A1* mRNA abundance.

**Figure 4:**
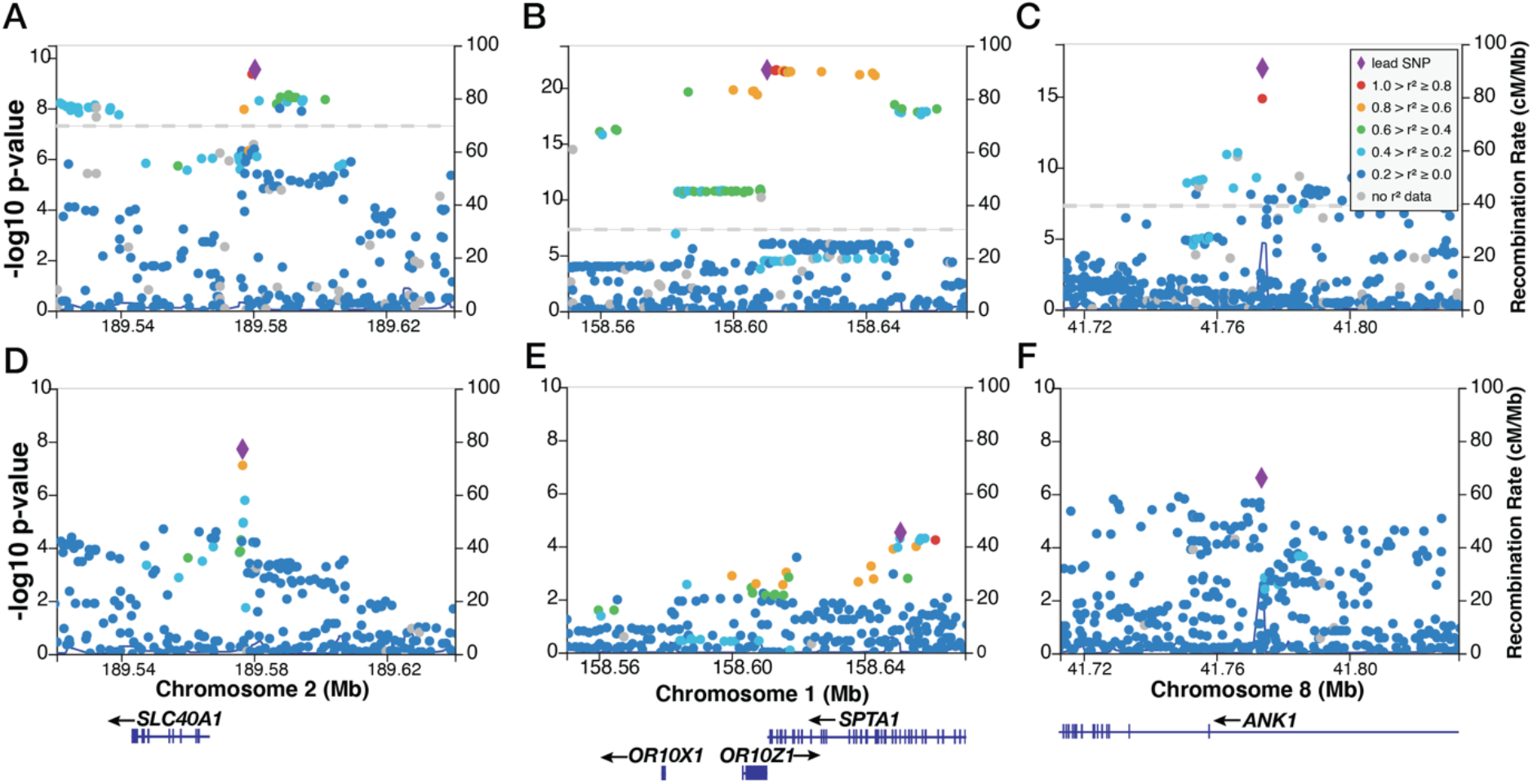
Common variants in *SLC40A1, SPTA1*, and *ANK1* loci are associated with spleen iron, and colocalize with cis-regulatory variation. **(A)** Fine-mapped locus near *SLC40A1* is associated with spleen iron (lead SNP: rs13008848). **(B)** Fine-mapped genome-wide significant locus near *SPTA1* is associated with spleen iron (lead SNP rs2479868). **(C)** Fine-mapped locus near *ANK1* is associated with spleen iron (lead SNP: 4373010). **(D)** Co-localization of cis-regulatory variation in the *SLC40A1* locus and spleen iron. Co-localization at this locus was observed in multiple tissues (posterior probability≥0.99 for each). Blood cis-eQTL data is shown. **(E)** Cis-regulatory variation in the *SPTA1* locus co-localizes with spleen iron. A signal was observed in multiple tissues but did not meet significance threshold in whole blood (posterior probability=0.72). **(F)** Cis regulatory variation at the *ANK1* locus co-localizes with spleen iron. Co-localization was observed in multiple tissues (posterior probability≥0.99). Grey dashed line indicates genome-wide significance (p=5e-8). For all eQTL signals, a threshold of FDR<5% was used in GTEx^38^. Linkage disequilibrium was calculated using 1000 Genomes Phase 3. Gene models are shown at bottom in GRCh38 coordinates.

The GWAS of spleen iron identified other loci relevant to iron homeostasis, including a locus on chromosome 9 (**Supplementary Figure 5B**). The lead SNP rs41276777[A] (beta=0.17, p=2.1e-9) occurred in the 5’ untranslated region of *PRPF4* and *CDC26* (**Supplementary Figure 5A**), and was associated with a regulatory locus affecting expression of both *PRPF4* and *CDC26* mRNA in whole blood and the spleen (**Supplementary Table 8**), suggesting bi-directional regulation of gene expression. This region colocalized with signals for serum ferritin, serum iron, and other blood iron traits (**Supplementary Table 7**), suggesting roles not only in spleen iron but body iron more broadly.

### Elevated spleen iron colocalizes with a splicing quantitative trait locus for *MS4A7* and monocyte traits

Our GWAS with spleen iron found a significant association at the *MS4A7* locus (lead variant: rs950802[A], beta=0.086, p=7.7e-26; **Table 2, Figure 5A**). rs950802[A] causes a synonymous mutation at Leu57 in the third exon of *MS4A7* and is also a variant in the first intron of *MS4A14*. This signal colocalized with a signal of serum ferritin^29^ (posterior probability≥0.99. Using splicing quantitative trait data^30^, we identified an alternative splicing event in *MS4A7*, which colocalized with spleen iron (**Figure 5B**). Based on an analysis of the open reading frame of *MS4A7*, this alternative splicing event was predicted to increase skipping of the second exon and therefore interrupt a conserved CD20-like transmembrane domain in *MS4A7* (**Figure 5C)**. We thus identified a novel association between *MS4A7* and spleen iron, and found a plausible molecular mechanism by which this variant disrupts *MS4A7* function.

**Figure 5:**
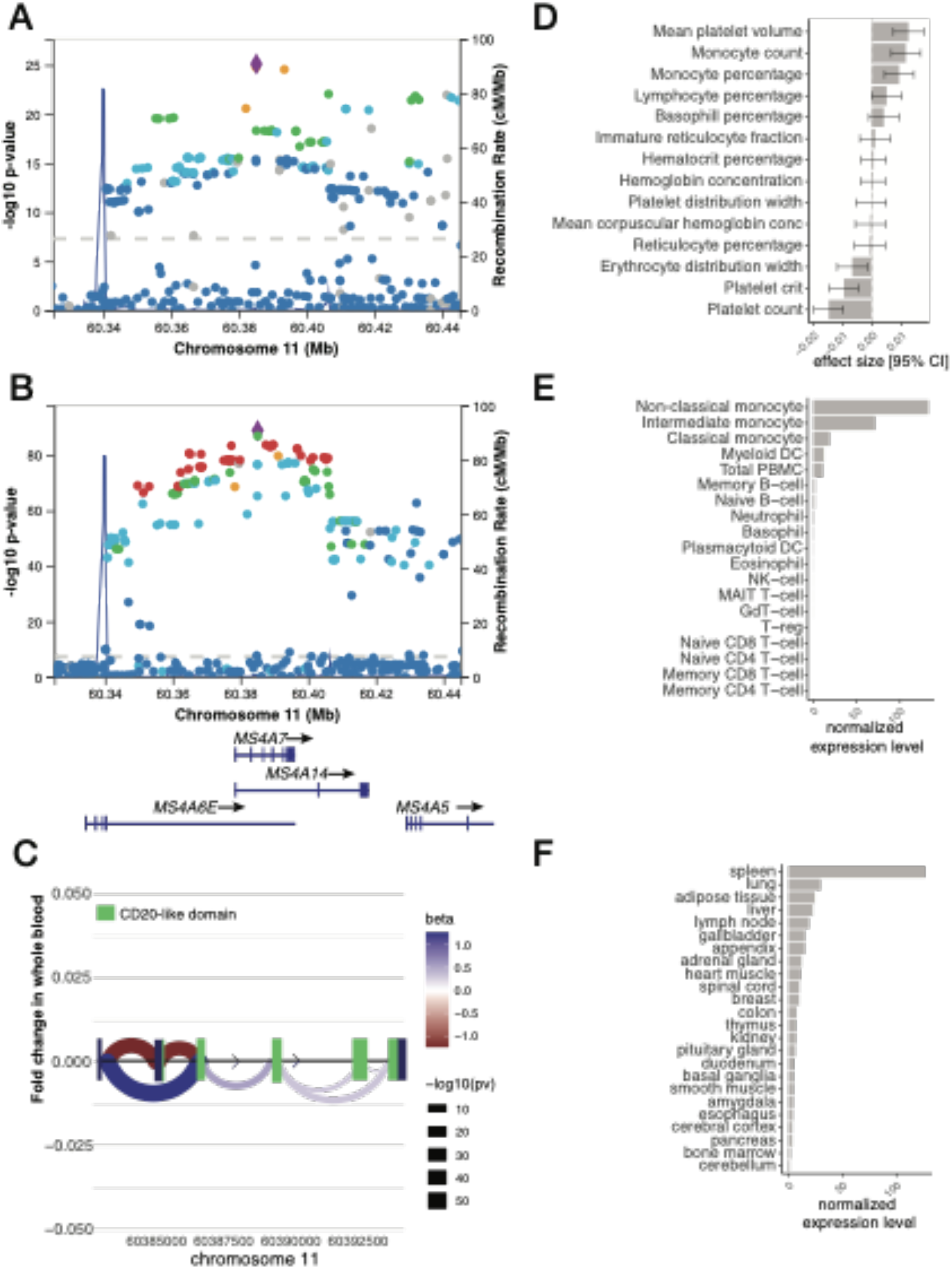
Characterization of the *MS4A7* locus. **(A)** The *MS4A7* locus on chromosome 11 is associated with an increase in spleen iron. Purple triangle displays the lead SNP rs950802[G] **(B)** This locus co-localizes with splicing quantitative trait locus of *MS4A7*, whole blood (posterior probability≥0.99). **(C)** This splicing quantitative trait locus is associated with exon skipping events in the *MS4A7* locus. Corresponding betas and p-value are shown. The *MS4A7* gene model is displayed with the conserved CD20-like domain shown in green. **(D)** Genetic associations with a panel of blood cell traits show associations with monocyte count, monocyte percentage and mean platelet volume, and negative associations with platelet count and platelet crit. **(E)** Expression of *MS4A7* was enriched in monocytes, including non-classical, intermediate, and classical forms. **(F)** Expression of *MS4A7* in human tissues displayed enrichment for lymphoid tissue, notably spleen and lymph nodes.

To explore the functional consequences of regulatory variation in *MS4A7*, we examined its association with hematological parameters in the UKBB, and observed a significant association with monocyte count and percentage, as well as platelet count and crit (**Figure 5D**). We replicated these findings in the Blood Cell Consortium^31^, confirming a positive association with monocyte count (beta=0.011, p=1.5e-8) and a negative association with platelet count (beta=-0.012, p=2.7e-10) in a European cohort (**Supplementary Table 10**). Further, we found enrichment of *MS4A7* mRNA in monocytes compared to sixteen other hematopoietic cell types^32^ (**Figure 5E**), and also higher expression of *MS4A7* mRNA in the spleen relative to other tissues (**Figure 5F**). A novel common allele at *MS4A7* was thus associated with elevated spleen iron and monocyte traits. *MS4A7* mRNA was enriched in the spleen and in monocytes, suggesting a role for this gene in iron recycling in the spleen.

### GWAS of spleen iron identifies novel common alleles in *SPTA1* and *ANK1* linked to increased gene expression and erythrocyte function

In addition to signals linked to iron homeostasis, the GWAS of spleen iron revealed associations in *SPTA1* and *ANK1*, encoding structural components of erythrocytes (**Table 2, Supplementary Figure 6**). The lead variant rs2479868[T] (beta=-0.083, p=2.5e-22) on chromosome 1 was located in the 3’ untranslated region of *SPTA1*, and the lead SNP rs4737010[A] (beta=-0.077, p = 1.0e-17) on chromosome 8 was the first intron of *ANK1* (**Figure 4B-C**). We tested for a shared signal with mRNA expression levels of *SPTA1* and *ANK1*, and observed co-localization with cis-regulatory variation for *ANK1* in multiple tissues (**Figure 4E-F**).

Variation at each locus was associated with increased mRNA expression and mean reticulocyte volume; and decreased mean corpuscular hemoglobin concentration, reticulocyte percentage, and spleen volume (**Figure 6**). The magnitude and directionality for the changes in red cell parameters associated with the lead SNPs at *SPTA1* and *ANK1* were similar, but a regression model testing for interaction between these two loci found that the effects are independent (beta=0.016, p=0.28). These signals did not co-localize with iron traits in other tissues such as serum or liver, suggesting specific effects to the spleen (**Supplementary Table 7**).

**Figure 6:**
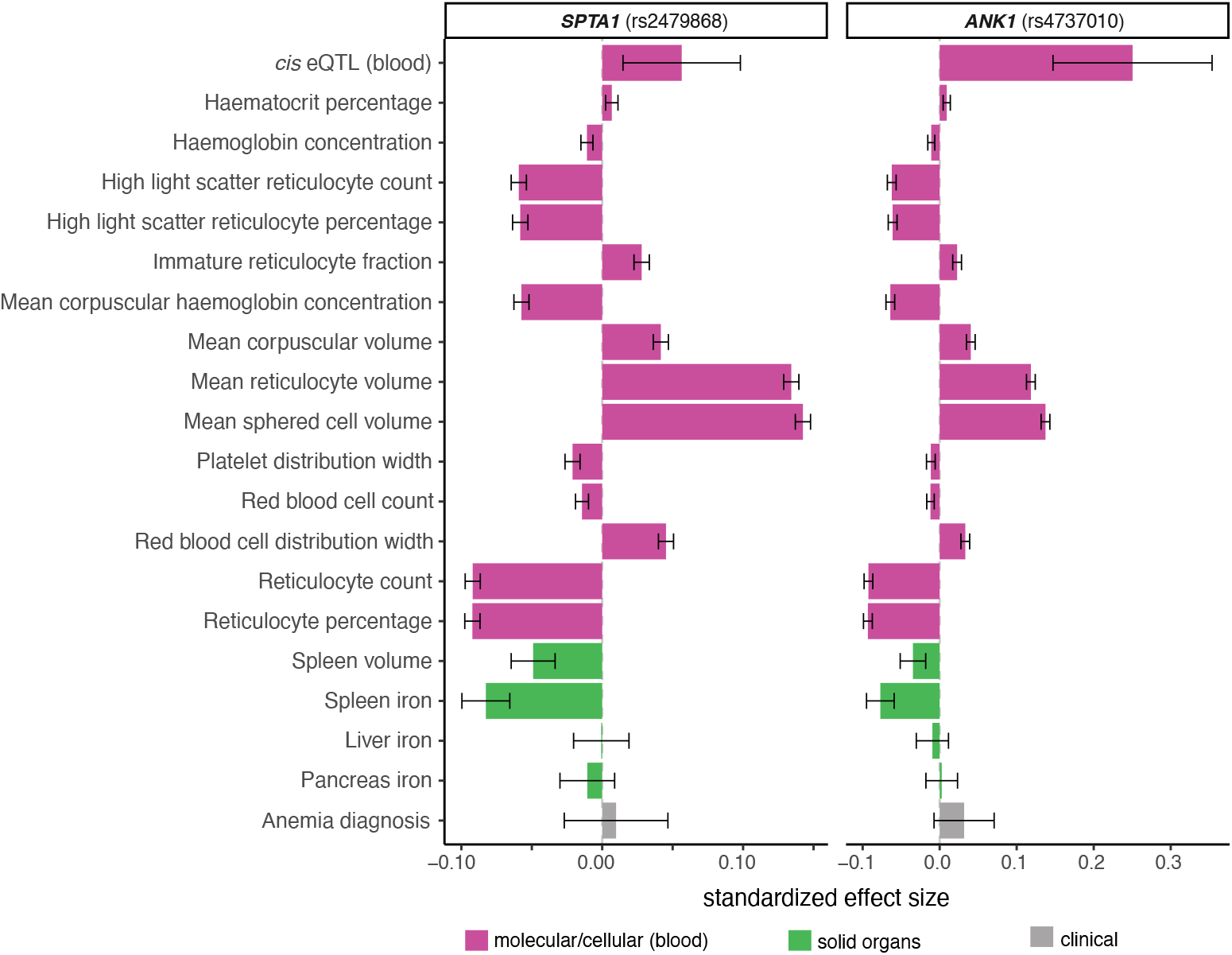
Associations of molecular, cellular, organ, and tissue traits with lead SNPs at *SPTA1* and *ANK1* loci indicate protective effects on red blood cell (erythrocyte) function. Both loci were associated with increased mRNA expression, beneficial effects on red blood cell parameters, decreases in spleen iron and spleen volume, and no effects on other measures of body iron or anemia diagnosis. Standardized effect sizes are shown. Error bars represent 95% confidence intervals.

Using data from the Blood Cell Consortium^31^, we replicated the associations and directions of effect between *SPTA1* and *ANK1* lead variants and mean corpuscular hemoglobin concentration and mean corpuscular volume. In a cohort of European ancestry, we replicated all findings (p<5e-10). Additionally, we observed associations for both loci and MCHC in East Asian and Hispanic/Latino ancestry cohorts (p<0.01) (**Supplementary Table 10**).

### Common alleles in *SPTA1* and *ANK1* show effects on erythrocytes opposite to effects observed with rare deleterious alleles and in hereditary spherocytosis

Given the known associations of both *SPTA1* and *ANK1* gene defects with hereditary spherocytosis, we analyzed rare variation (minor allele frequency <0.001) predicted to cause loss-of-function. Starting with 167,246 exomes of European ancestry, we conducted rare variant association studies for reticulocyte percentage and volume and identified one significant gene, *SPTA1*, in both studies (p_SKAT-O_< 1e-24; **Supplementary Figure 7**). Performing a scan of the same loss-of-function rare variation in *SPTA1* across a hematology panel, we recapitulated clinical hallmarks of spherocytosis: *SPTA1* loss-of-function was significantly associated with increased reticulocyte percentage and increased MCHC, increased bilirubin, decreased mean spherical cell volume and decreased MRV (**Supplementary Figure 7)**. We did not observe a significant association with spleen iron in the imaging subcohort for either putative loss-of-function or deleterious missense variation, perhaps due to reduced statistical power of the imaging subcohort (n=18,420) compared to the exome cohort (n=167,246) (**Supplementary Figure 8**).

Finally, we genetically identified HS in the UKBB exome cohort using clinical assertions of pathogenicity and predicted high confidence loss-of-function in one of six HS genes, and estimated prevalence to be 1:389 [95% CI 1:427 - 1:354]. We asked whether the common alleles found via GWAS could modify the effects of rare deleterious alleles for hematology parameters relevant for HS, including mean reticulocyte volume and percentage. We estimated that carrying either of the *SPTA1* or *ANK1* lead GWAS SNPs skewed erythrocyte parameters toward beneficial effects in deleterious allele carriers and non-carriers, suggesting that the common GWAS alleles could modify the effects of the rare deleterious alleles (**Supplementary Figure 10**).

## Discussion

Most studies of spleen iron have not included healthy volunteers, or methods that reliably assess organ iron^15,33–35^ or only provided qualitative histological grading^35,36^ (**Supplementary Table 11**).^14,17–19^ In this study, we combined deep learning algorithms and efficient image processing to quantify spleen iron in 41,764 participants of the UKBB. This enabled us to estimate the first reference range for spleen iron in a large, unselected population.

Spleen iron was higher in men, and our analysis of pre-and post-menopausal women is consistent with prior reports of iron stores in other tissues.^37^ Spleen iron increased with age, is associated with red meat intake and inversely associated with alcohol consumption, extending prior observations in the liver.^38^-_39,40_Further, spleen iron was only modestly associated with measures of iron in other tissues. Unlike other iron-rich organs such as the liver, we discovered that spleen iron was associated with indicators of reticulocytosis and reticulocyte turnover.

Our genome-wide association study of spleen iron identifies regulatory loci in *SPTA1* and *ANK1*, which when combined with other evidence, suggests a model of low splenic turnover due to relatively large, long-lived erythrocytes. These alleles were associated with decreased spleen iron and increased mRNA expression of their respective *cis* gene, as well as larger reticulocyte volume, and reduced measures of reticulocytosis (**Figure 6**). Since the spleen is the major route of erythrocyte clearance and iron salvage, lower levels of reticulocytosis would be expected to result in lower spleen iron at steady state. Neither variant was associated with anemia, suggesting that they are not pathogenic. This reduced-turnover model also predicts that iron levels in other organs not involved in erythrocyte clearance (such as the pancreas) would be unaffected by these two loci, consistent with the data.

As genetic modifiers, these alleles in *SPTA1* and *ANK1*, which segregate frequently across global populations, may explain the variable penetrance and expressivity observed in hereditary spherocytosis (HS). In a recent study of patients with identified HS mutations, 64% involved *SPTA1* or *ANK1*, and the investigators observed multiple HS families with broad phenotypic variability, including a compelling example of dizygotic twins sharing the same pathogenic *ANK1* mutation presenting with mild disease in one case and severe disease requiring splenectomy in the other^11^. The variation could not be explained, leading to speculation that yet-unknown genetic factors may be contributing. We were able to identify the hallmarks of HS through rare loss-of-function variation in *SPTA1*, even within a cohort not ascertained for hemolytic anemias. Our analysis suggested that common genetic variants may modify the effects of rare deleterious alleles. To further support this, we note that both common alleles deviated from Hardy Weinberg equilibrium (p= 9.0e-8 for *SPTA1* lead SNP, p=7.28e-17 for *ANK1* lead SNP), suggesting that they could be under positive selection. The expression-increasing variants identified here may help to explain the heterogeneity that has been observed in HS patients.

In addition to genes specific to red cell turnover in the spleen, our genetic study of spleen iron also pointed to known and novel regulators of the human body’s iron economy. We characterized a spleen iron signal at *MS4A7*, which belongs to the CD20 family of membrane proteins, which are expressed within the hematopoietic lineage and largely uncharacterized.^42,43^ Here, we linked a splicing quantitative trait locus in *MS4A7* to spleen iron, macrophage abundance, and even found that this gene was enriched in the spleen and in macrophages. It is possible that excess circulating monocytes can be recruited to provide an expanded reservoir in the spleen, contributing to the body’s iron economy in addition to splenic macrophages^44^. Our findings regarding *MS4A7* potentially illuminate additional details of splenic erythrocyte clearance mechanisms. For example, the specific functional roles played by macrophages and monocytes in the splenic red pulp remain incompletely understood.^44,9^

This study has limitations. First, as the only study to quantify spleen iron in a large cohort, no replication cohort is available, though we are able to replicate all our findings on relevant blood cell traits in independent cohorts. Second, while we found limited evidence in the UK Biobank that spleen iron varies by ethnicity (**Supplementary Figures 3-4**), additional imaging studies are warranted to quantify spleen iron across populations. Third, experimental studies in the hematopoietic lineage of a model system will be needed to test the functional consequences of the regulatory variation observed here. Fourth, while we genetically identified HS in the UKBB and showed that the common alleles in *SPTA1* and *ANK1* act as genetic modifiers of erythrocyte parameters, additional clinical validation is needed to substantiate these findings in an HS patient cohort.

In summary, we have quantified spleen iron by repurposing liver MRI acquisitions, maximising use of the data at no extra cost to scanning or participant time. Our findings suggest that steady-state levels of spleen iron are sensitive to alterations in erythrocyte structure affecting cell turnover, as well as alterations in iron transport by macrophages. We identified common regulatory variation in HS genes showing effects on erythrocytes that are opposite of the effects observed in HS patients. Imaging-derived spleen iron is thus a novel quantitative biomarker that is tractable for genetic analysis, and has potential to contribute to future clinical characterization of hemolytic anemias.

## Methods

### Image Analysis

Image processing and segmentation pipelines are described in Liu et al.^26^ We extracted 2D masks from the 3D spleen segmentations at their intersection with the liver acquisition (**Figure 1A**).^45^ We applied one-pixel erosion before computing the median value within that mask. We excluded 2D masks that had < 1% of 3D volume or < 20 voxels. Additional methods are described in the Supplement.

### Epidemiological modelling of spleen iron risk factors

Associations of spleen iron with age, genetic sex, and self-reported ethnicity were performed in R v3.6.3. using the linear and logistic models for spleen iron as a quantitative or binary trait, respectively, after adjusting for covariates including imaging center, date and time.

### Phenome-wide association study

We generated a list of variables derived from raw data used PHESANT^46^, and removed procedural metrics (e.g. measurement date), duplicates, and raw measures, resulting in 1,824 traits (**Supplementary Table 2**). We used PheWAS^47^ to combine ICD10 codes (Field 41270) into distinct phenotype codes or phecodes (**Supplementary Table 3**). We performed linear (quantitative traits) or logistic regression (binary traits) on spleen iron, adjusting for imaging center, date, scan time, age, sex, BMI, height, and ethnicity.

### Genome-wide association study and replication analysis

We conducted a GWAS in n=35324 participants using methods as previously described.^48^ Additional methods are described in the Supplement. Summary statistics from the Blood Cell Consortium^31^ were downloaded from http://www.mhi-humangenetics.org/en/resources/ and harmonized using dbSNP build 151 (GRCh37). Using relevant blood cell traits, we tested for replication using all available ancestries: Europeans, East Asians, African-Americans, and Hispanic/Latinos.

### Conditional analysis and fine-mapping

We performed conditional analysis using GCTA^49^, considering variants within 500kb of an index variant. We constructed a reference LD panel of 5,000 randomly selected, unrelated European UKBB participants^24^. We excluded the major histocompatibility complex region due to the complexity of LD structure at this locus (GRCh37::6:28,477,797-33,448,354; see https://www.ncbi.nlm.nih.gov/grc/human/regions/MHC). For each locus, we considered variants with locus-wide evidence of association (p_joint_<10^−6^) to be conditionally independent. We followed an iterative procedure to determine credible sets of causal variants with 95% coverage.^48^

### Colocalization studies

For gene expression studies, we used summary statistics from GTEx v8^50^. For disease and quantitative trait studies, we used UKBB summary statistics of phecodes^51^, normalized quantitative traits (http://www.nealelab.is/blog/2017/7/19/rapid-gwas-of-thousands-of-phenotypes-for-337000-samples-in-the-uk-biobank). We selected phenotypes with p<5×10^−8^ within 500kb of the index variant. We performed colocalization analysis using coloc^52^ with default priors and considered variants within 500kb of the index variant. We considerd two genetic signals to have strong evidence of colocalization if PP3+PP4≥0.99 and PP4/PP3≥5.^53^

## Data Availability

Model weights for spleen segmentation are available via Github (www.github.com/calico-ukbb-mri-sseg) and code for resampling at www.github.com/recoh/pipeline. All summary statistics will be made available from the GWAS Catalog (www.ebi.ac.uk/gwas) upon publication. All derived data will be made available from the UK Biobank (www.ukbiobank.ac.uk).

## Acknowledgments

The authors would like to thank David Botstein, Marcia Paddock, Amoolya Singh, Kevin Wright, and Bert van der Zwaag for helpful feedback and discussion. This work was made possible by the UK Biobank, including staff, funders, and study volunteers. This research has been conducted using the UKBB Resource under Application Number 44584 and was funded by Calico Life Sciences LLC.

## Author Contributions

NB, MC, EPS, JDB, ELT designed the study. NB, BW, and YL implemented the image processing methods. MC performed the data processing. EPS and MC performed genetic analysis. ELT, EPS, RLC, MC and NB drafted the manuscript. All authors edited, read, and approved the manuscript.

