## Supplementary Information for "Analysis of MRI-derived spleen iron in the UK Biobank identifies genetic variation linked to iron homeostasis and erythrocyte morphology"

### Supplementary methods

#### Image Acquisition

We used the UK Biobank (UKBB) liver single-slice multi-echo MRI and the neck-to-knee Dixon MRI[25] in 44,265 subjects. For the first approximately 10,000 subjects, a gradient-echo (GRE) sequence was used, and subsequently an IDEAL[60] sequence was used (1,364 participants had both and contained the spleen). The liver slice covers the abdominal width generally including the spleen due to its position relative to the liver. **Figure 1** shows a coronal view of the neck-to-knee Dixon and the location of the liver 2D slice in red **(A)**, the estimated iron content for the liver and spleen in the multi-echo slice **(B)**.

#### GRE and IDEAL acquisitions

Using the 1,364 participants with both the IDEAL and GRE acquisitions, we compared the R2* and PDFF measurements (**Supplementary Figure 2**). Spleen PDFF in the GRE and IDEAL acquisitions were not correlated, supporting the idea that measurements of spleen PDFF via MRI are the result of noise reconstruction.[61] We therefore did not perform subsequent analysis on the PDFF measurements but included them for completeness as a negative result. Spleen R2* in the GRE and IDEAL acquisitions were strongly correlated ($R^2$=0.86).

To combine data from the GRE and IDEAL acquisitions, we first ranked the measurements within each modality. Where individuals had both measurements, we averaged the ranks. We then applied an inverse normal transformation and performed subsequent analyses using these residuals.

In order to report population-level iron concentration values while accounting for systematic differences between the acquisitions, for individuals with only GRE R2* measures, we applied a linear transformation. This transformation was derived by fitting a linear model to the 1,364 individuals with spleen R2* measurements for both IDEAL and GRE modalities:

$$R2^*_{IDEAL} = -1.276 + 1.2366 * R2^*_{GRE}$$

We then transformed R2* to iron (mg/g) as previously described.[48,39]

#### Validation of opportunistic resampling strategy

In order to validate our opportunistic resampling strategy used to obtain 2D spleen masks, we used direct neural network-based liver 2D segmentations from a previous study[26] and compared those to opportunistic resampling of the liver (**Supplementary Figure 1).** For n=38,400, the results of opportunistic resampling against dedicated 2D liver segmentation shows a correlation of 0.98 for proton density fat fraction (PDFF) and 0.94 for R2*.

#### MRI-derived PDFF of spleen is noise

While measurement of spleen fat was possible, with measured values ranging between 0 and 4.98% (**Supplementary Table 1**), close scrutiny of these measurements for n=1,364 subjects who had two different acquisitions covering the same anatomy indicate a level of background noise (**Supplementary Figure 2B**; $R^2$=0.01) leading us to agree with Hong et al.[61], that MRI measured spleen fat is at the level of background noise and cannot be meaningfully quantified. By contrast, spleen iron was reproducibly measured from both GRE and IDEAL (**Supplementary Figure 2A**; $R^2$=0.86). Therefore, we focused subsequent analyses on spleen iron. Spleen iron is shown in **Table 1** as mean ± standard deviation.

##### Identification of GRE acquisitions with high iron content

Through the quality control process, it has come to our attention that the GRE echo times in the UKBB abdominal MRI protocol are not short enough to quantify iron when it is exceeding a threshold (approximately 4 mg/g). High iron concentrations have a superparamagnetic effect that distorts the local magnetic field resulting in a faster decay of transverse magnetization.[41] Values beyond this threshold are not detected in GRE and would be falsely assigned a low value unless flagged up. However, this issue may be accurately identified from the magnitude data across echo times. Using the 2D organ segmentations, we calculated the average signal intensity for each echo time. When the decay of these values had a drastic signal dropoff and the average values of the longest echo time magnitude images displayed no decay, we estimated that the iron is mischaracterized. We assigned a maximum iron value of 4 mg/g to those participants.

#### Genome-wide association study

We performed GWAS as described[49], using UKBB imputed genotypes[24] version 3, excluding single nucleotide polymorphisms (SNPs) with minor allele frequency <1% and imputation quality <0.9. We excluded non-European participants, participants exhibiting sex chromosome aneuploidy, participants with a discrepancy between genetic and self-reported sex, heterozygosity and missingness outliers, and genotype call rate outliers.[24] 9911384 SNPs passed our QC thresholds and were included in the study. We used BOLT-LMM[50] v2.3.2 to conduct the genetic association study. We included age at imaging, age squared, sex, imaging centre, scan date and time, and genotyping batch as fixed-effect covariates, and genetic relatedness derived from genotyped SNPs as a random effect to control for population structure and relatedness, and normalized the outcome variable using inverse rank normalization. In the genetic association study, we found no evidence for global inflation of test statistics ($\lambda_{gc}$=1.035; LD score regression intercept 1.027 [s.e. 0.0072])

#### Heritability estimates

We estimated the heritability of each trait using the restricted maximum likelihood method[51], as implemented in BOLT-LMM.

#### Genetic correlation

We computed genetic correlation using bivariate LDSC.[52] We computed the genetic correlation between spleen iron and 288 complex traits with a heritability of at least 5% from the Neale Lab http://www.nealelab.is/uk-biobank/, plus organ iron measurements[26], and blood iron biomarkers.[29] We used a Bonferroni-corrected p-value of 1.7e-4 as the significance threshold.

#### Exome sequence quality control

We performed quality control of n=200,643 whole exomes from the UKBB using our custom "FE-plus" pipeline. Raw genotype calls were filtered genotype-level quality metrics to identify quality outliers for a given site, and remove poor-quality individual-level genotypes. Similarly to the FE pipeline[62], we removed genotypes below a minimum read depth (for SNPs: 7 and for indels: 10), and genotypes below a minimum Phred-scaled genotype quality of 20. We removed genotypes where minor allele allelic balance < 0.15 for SNPs and 0.2 for indels. Supplementing FE filters with additional filters, we performed per-SNP QC, requiring the average genotype quality to be at least 30 and per-SNP depth of coverage to be at least 15, to filter out badly captured sites. Additionally, we removed variants with genotype missingness > 10% or that deviated meaningfully from Hardy Weinberg equilibrium in a European ancestry cohort (HWE p< 1e-10). Of 17,981,897 total variants, 13,907,865 variants passed QC in the European exome cohort with MRI data (n=18,240).

#### Filtering self-reported ancestry for outliers

Self-reported ethnicity continental ancestry (European, South Asian, African, and East Asian) was obtained from the UKBB (field 21000) and principal component analysis performed using common (minor allele frequency >=0.05), independent ($r^2$<0.2), autosomal markers, in Hardy Weinberg equilibrium (p>1e-10), excluding regions of long-range linkage disequilibrium. FlashPCA v2.1 was used to calculate the first 10 principal components (PCs). Centroid distance was calculated by subtracting the population PC mean from the individual's PC, squaring, and dividing by the variance for that PC. Ancestry outliers were identified as extreme values on a histogram of centroid distance for PCs 1-3.

#### Exome variant annotation

We performed annotation using VEPv100, LOFTEE[60], CADD[61] and ClinVar (https://www.ncbi.nlm.nih.gov/clinvar/, downloaded on September 27, 2020) with a custom pipeline to select variants meeting high-confidence loss-of-function criteria, further filtered for rare variants (defined as cohort-specific minor allele frequency <0.001), see Supplemental methods for QC. Of 13,907,497 total variants passing quality control, we subset 286,456 high-confidence, rare loss-of-function variants, 2,919,962 rare missense variants (CADD sore >=20), and 13,705 rare clinical pathogenic variants in 19,992 European ancestry samples for further analysis.

#### Rare variant regression study

We performed rare variant burden and SKAT testing as implemented in SAIGE-GENE, using a mixed-effects model.[62] A kinship matrix was built in SAIGE from a filtered set of 354,878 genotyped variants ($r^2$<0.2, minor allele frequency >0.05, Hardy-Weinberg p-value >1e-10, excluding known regions of long-range linkage disequilibrium). The linear mixed model regression equation was as follows:

$$y_i = \alpha + X_{1,i}\beta_1 + X_{2,i}\beta_2 + X_{3,i}\beta_3 + X_{4,i}\beta_4 + X_{5,i}\beta_5 + X_{6,i} + X_{7,i}\beta_7 + X_{8,i}\beta_8 + \sum_{j=1}^{5} PC_{ij}\beta_j + G_i\beta + b_i + \epsilon_i$$

In the model, $y_i$ is inverse-rank normalized spleen iron, $X_1$ is age at imaging visit, $X_2$ is age$^2$, $X_3$ is chromosomally determined sex expressed as a binary indicator variable, $X_4$ and $X_5$ represent the categorical variable of study center as two dummy variables variables, $X_7$ is standardized scan date, $X_8$ is standardized scan time, $PC_j$ represents the first five principal components of European genetic ancestry, and $G_i$ represents the allele counts (0,1,2) for q variants in each gene to test. We then performed SKAT and burden tests in SAIGE-GENE[62] and reported both the p-value from SKAT-O, which is a linear combination of burden and SKAT: $Q_{SKAT-O} = (1-\rho)Q_{SKAT} + \rho Q_{burden}$where $\rho$ is estimated as previously described.[62] To inform directionality of effect, we reported the betas from the burden test. To avoid unstable results at low sample size, we calculated cumulative minor allele count and thresholded at >=5 minor alleles per gene, including singletons and doubletons. Genomic inflation factor was calculated using the gap package and summary statistics were visualized in R using the ggplot package.

#### Identification of clinical carriers of *HFE,* hemoglobin, and *G6PD* alleles

A version of ClinVar was downloaded on February 1, 2021, from https://ftp.ncbi.nlm.nih.gov/pub/clinvar/vcf_GRCh37/. Pathogenic, likely pathogenic, and pathogenic/likely pathogenic variants were selected with a level of evidence of either a single submission with criteria provided, multiple submitters agree, or reviewed by an expert panel (there are no variants in hemoglobin genes with a practice guideline). Genotypes were calculated using PLINK v1.90 and R 3.6.3 to assess compound genotypes.

#### Genetic identification of hereditary spherocytosis alleles

From the exome cohort filtered for European ancestry (n=167,246), we annotated variants by clinical assertion as pathogenic according to ClinVar, downloaded from https://ftp.ncbi.nlm.nih.gov/pub/clinvar/ on September 27, 2020, and called predicted loss-of-function alleles using LOFTEE[63]). HS alleles were defined as clinical pathogenic variants or high-confidence putative loss-of-function alleles in one of six genes (*SPTA1, SPTB, SLC4A1, ANK1, EPB41,* or *EPB42).*

### Supplementary results

#### Spleen iron varies by ethnicity

Spleen iron varied by self-reported ethnicity, even after pruning ancestry outliers and adjusting for age, sex, study center, scan date, and time ($p_{Kruskal\text{-}Wallis}$=2.1e-20; Methods, **Supplementary Figure 4**). Even after removal of the British group (n=32,414) which was considerably larger than the other groups, there were differences in group means ($p_{Kruskal\text{-}Wallis}$=1.1e-17). We performed a similar analysis using liver iron, adjusted for the same covariates, and observed differences in group means ($p_{Kruskal\text{-}Wallis}$=2.1e-4; **Supplementary Figure 4**). We note that the non-European groups were of small sample size in this study, and larger sample sizes are needed to confirm these observations.

#### HFE carriers and compound heterozygotes have elevated spleen iron as compared with non-carriers

HH is a recessive Mendelian disorder characterized by accumulation of iron in the liver, blood and other tissues and is most commonly caused by missense variants in the *HFE* gene, which regulates iron uptake into hepatocytes in the liver. We examined two missense variants, HFEp.Cys282Tyr and HFEp.His63Asp, both of which segregate in the UKBB at high frequency (HFEp.Cys282Tyr: minor allele frequency (MAF) = 0.146 [95% CI 0.145 - 0.146] HFEp.His63Asp: MAF = 0.073 [95% CI 0.073 - .074]). While we observed elevated liver iron in C282Y and H63D carriers, compound heterozygotes, and homozygotes (**Supplementary Figure 9A**), we did not observe any effects of HH carrier status on spleen iron, after adjustment for covariates (**Supplementary Figure 9B**).

Thalassemia is a blood disorder characterized by low levels of hemoglobin. Although there was only one participant with a confirmed diagnosis in the imaging cohort, we identified putative carriers of mutations in globin genes characterized as pathogenic or likely pathogenic in ClinVar. We detected 345 mutations in globin genes (*HBA, HBB, HBC, HBD, HBE, HBF, HBH,* and *HBS*) asserted as pathogenic or likely pathogenic in ClinVar. Of those, only three variants in HBA and 20 variants in *HBB* segregated within the entire UKBB. All of these segregating variants were extremely rare, with the most common variant being rs33946267[C] in HBB, with a minor allele frequency of MAF=3.2e-4 in the entire UKBB. None of these carriers overlapped with the imaging cohort, precluding further analysis of thalassemia carriers.

Glucose-6-phosphate dehydrogenase (G6PD) deficiency is a genetic metabolic abnormality particularly in males, caused by deficiency of the enzyme encoded by *G6PD* which is associated with RBC dysfunction. We identified 15 pathogenic variants underlying G6PD deficiency. Here, three variants did segregate in the imaging subcohort, most commonly rs1050828[C], with a minor allele frequency of 1.2e-3. Within the imaging cohort, there were 19 males with one of three *G6PD* pathogenic mutations. We compared spleen iron in *G6PD* carriers to non-carriers in males, but did not observe a significant difference at this sample size (**Supplementary Figure 5**).

#### Supplementary Tables

**Supplementary Table 1:** Values for spleen and liver iron, PDFF, and R2* divided into GRE and IDEAL acquisitions in the UK Biobank.

**Supplementary Table 2:** Spleen iron concentrations in the UK Biobank, stratified by males and females.

**Supplementary Table 3**: Results from phenome-wide association study of spleen iron with 2,346 traits including medical history, lifestyle factors, and self-reported questionnaires.

**Supplementary Table 4**: Results form phenome-wide association study of spleen iron with 857 disease diagnosis codes.

**Supplementary Table 5**: Genetic correlations between spleen iron and 288 traits in the UK Biobank.

**Supplementary Table 6**: Genetic correlations between spleen iron and six previously published blood and organ iron traits: ferritin, serum iron, transferrin saturation (TSAT), and total iron binding capacity (TIBC)[29], and pancreas and liver iron[26].

**Supplementary Table 7**: Colocalization test results for genome-wide significant loci from the spleen iron GWAS with loci from six previously published blood and organ iron traits.

**Supplementary Table 8**: Colocalization test results for genome-wide significant loci from the spleen iron GWAS with expression quantitative trait loci (eQTLs) from 56 tissues of the GTex consortium (version 8).

**Supplementary Table 9**: Colocalization test results from genome-wide significant loci from the spleen iron study with hematological assays.

**Supplementary Table 10:** Replication analysis of *SPTA1* and *ANK1* associations with red cell parameters in an independent cohort of the Blood Cell Consortium, representing East Asian, African-American, Hispanic/Latino and European populations.

**Supplementary Table 11. Comparative data from spleen iron measured by MRI in healthy volunteers and patient groups**. T2* published values were converted into R2* values using the equation R2* = 1000/T2*. R2* published values were converted into [FE} in mg/g[48]. *Mean age only provided for mixed cohort, ↑Mean values calculated from individual patient values provided in the publication. MRI measurement of spleen iron may be expressed as signal intensity ratio based on T2w or T2*w imaging, T2, R2, T2* or R2*[33] or converted into values in mg/g using formulas derived from liver studies[48,64]. Older methods using changes in T2 relaxation are not as sensitive for mild or moderate spleen iron overload.[35] The upper range of T2* measures are higher than those measured using R2*, limiting direct comparison[34], furthermore, the relationship between R2 and R2* is different in the liver and spleen therefore R2 measurements may underestimate spleen iron[15], as such, they have not been included in this comparison.

#### Supplementary Figures

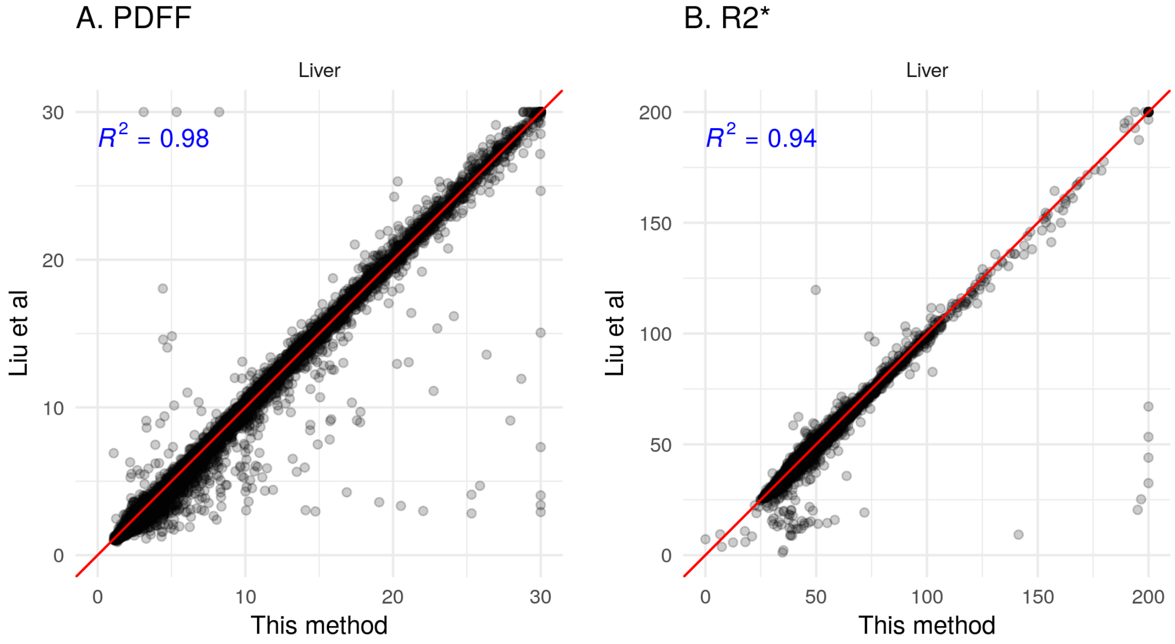


**Supplementary Figure 1: Scatter plot comparing direct neural network-based segmentation[26] in a dedicated 2D slice (y-axis) with opportunistic resampling[47] from the 3D (Dixon) acquisition (x-axis)** A) median liver PDFF (in %) and (B) median liver R2* (in $s^{-1}$) (n=38,400).

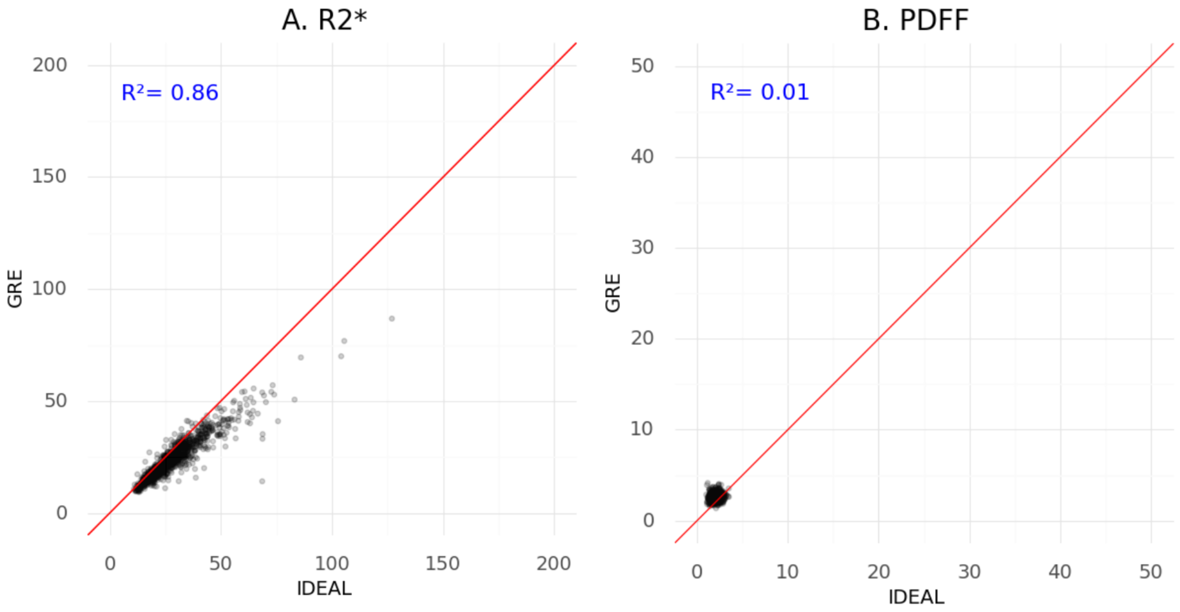


**Supplementary Figure 2. Relationship between parameters measured in the spleen in n=1,364 UKBB subjects with both GRE and IDEAL single-slice multi-echo acquisitions**. Scatter plots for median R2* (A) measurements (in $s^{-1}$) and PDFF in % (B). Pearson's correlation between the measures is given in blue.

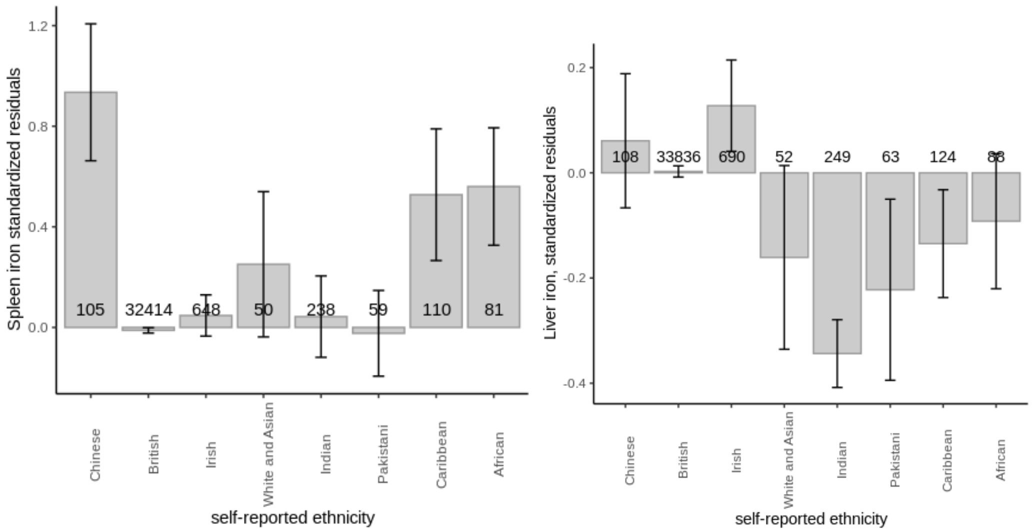


**Supplementary Figure 3: Spleen iron varies by self-reported ethnicity.** (A) Spleen iron displayed as standardized residuals after adjustment for age, sex, study center, scan date and time. (B) Liver iron content displayed as standardized residuals after adjustment for the same covariates. Only groups with >50 individuals with imaging phenotypes are shown. 95% CIs are shown.

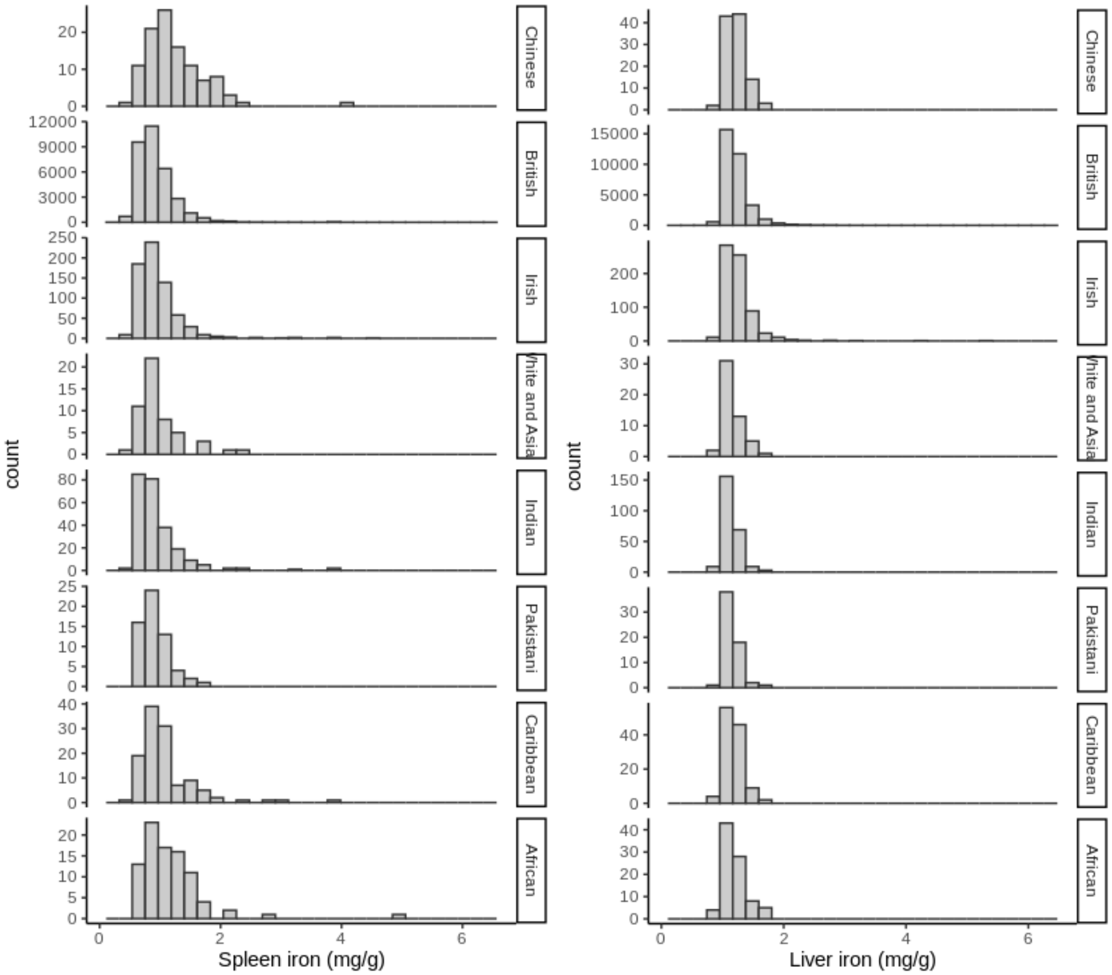


**Supplementary Figure 4**: **Spleen iron (in mg/g) varies by self-reported ethnicity but with a limited sample size for non-European groups**. Left panel: Histogram of spleen iron in mg/g by self-report ethnicity. Right panel: Histogram of liver iron content in mg/g by self-report ethnicity.

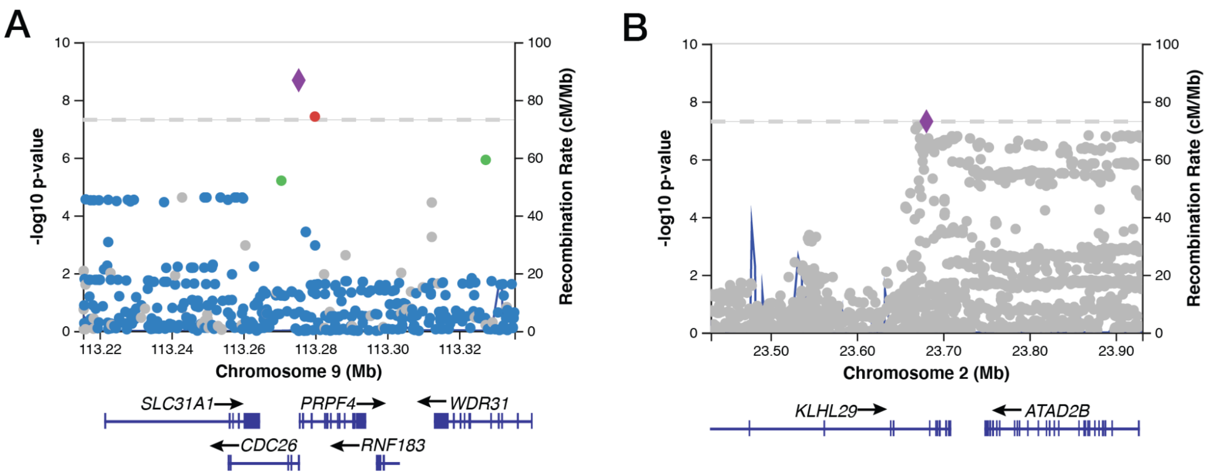


**Supplementary Figure 5: Lead SNPs associated with spleen iron in the *PRPF4/CDC26* and *KLHL29* loci after statistical fine-mapping.** (A) The lead SNP in the *PRPF4/CDC26* locus on chromosome 9 is rs41276777[A]. (B) The lead SNP in the *KLHL29* locus on chromosome 2 is rs115697725[G].

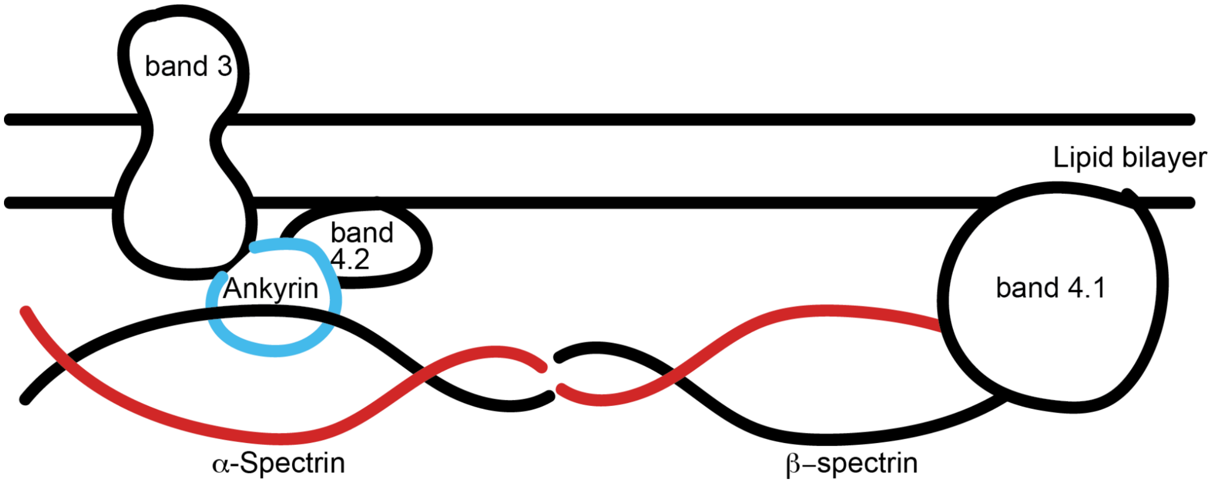


**Supplementary Figure 6: Simplified cartoon of a cross-section of an red blood cell membrane cytoskeleton.** Cytoskeletal protein ankyrin (encoded by the *ANK1* gene) is shown in blue, and alpha-spectrin (encoded by *SPTA1*) is shown in red. Alpha and beta spectrins contain antin-binding domains and associate to form anti-parallel heterodimers which interact to form tetramers. Ankyrins anchor anion exchangers in the lipid bilayer to the spectrin filaments. The anion exchanger band 3, as well as cytoskeletal proteins band 4.1, and band 4.2, are depicted for reference. Adapted from Delaunay et al.[6]

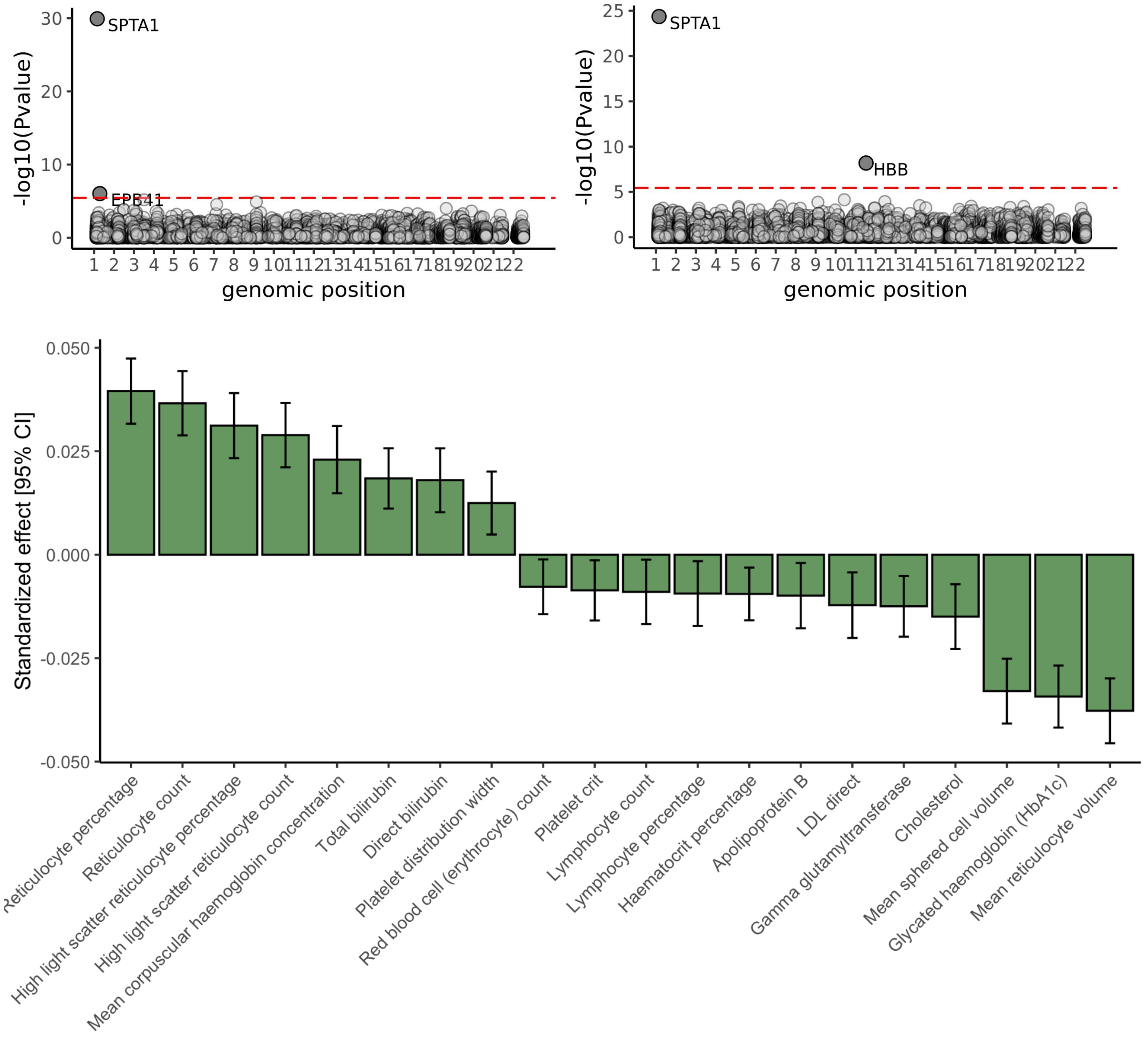


**Supplementary Figure 7: Predicted loss-of-function variation in *SPTA1* is associated with RBC biomarkers, platelet biomarkers, bilirubin, cholesterol, and glycated hemoglobin (HbA1c) in 167,243 exomes via rare variant association study (RVAS). Top left**: Exome-wide association study (ExWAS) of reticulocyte percentage identifies two components of red blood cell membrane architecture: *SPTA1* (encoding alpha-spectrin) and *EPB41* (encoding band 4.1). **Top right:** ExWAS of mean reticulocyte volume identifies *SPTA1* and *HBB*, encoding beta hemoglobin Bonferroni significance is shown with the dashed red line. **Bottom:** ExWAS across a panel of 31 quantitative hematological traits for rare coding mutations in *SPTA1* recapitulated signatures of hereditary spherocytosis, including increased reticulocyte percentage, mean corpuscular hemoglobin concentration, bilirubin; and decreased mean sphered cell volume and mean reticulocyte volume.

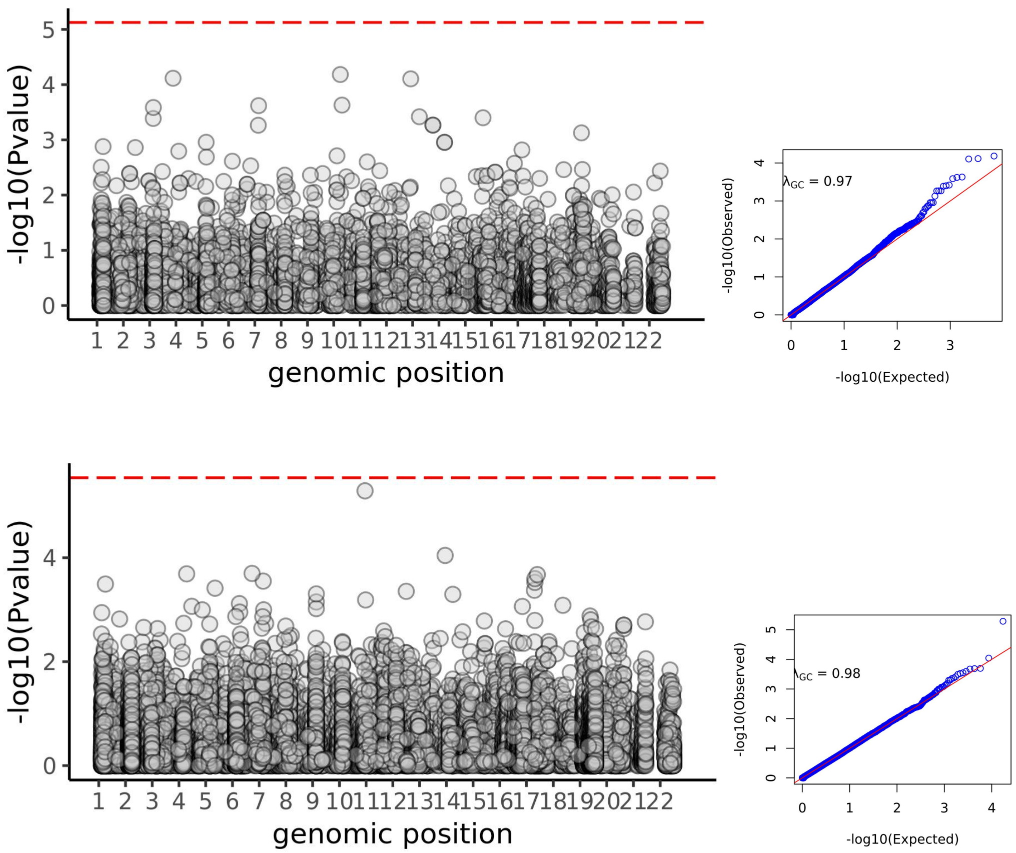


**Supplementary Figure 8: Rare variant burden testing to identify associations with spleen iron**. **Top panel:** Rare variant burden testing of loss-of-function variation in 18,240 exomes. 286,546 variants in 6,686 genes (minor allele count >=5) were tested. QQ plot is shown at right. **Bottom panel:** Rare variant burden testing of predicted deleterious missense variation. 2,919,962 variants in 17,298 genes (minor allele count >=5) were tested. QQ plot is shown at right.

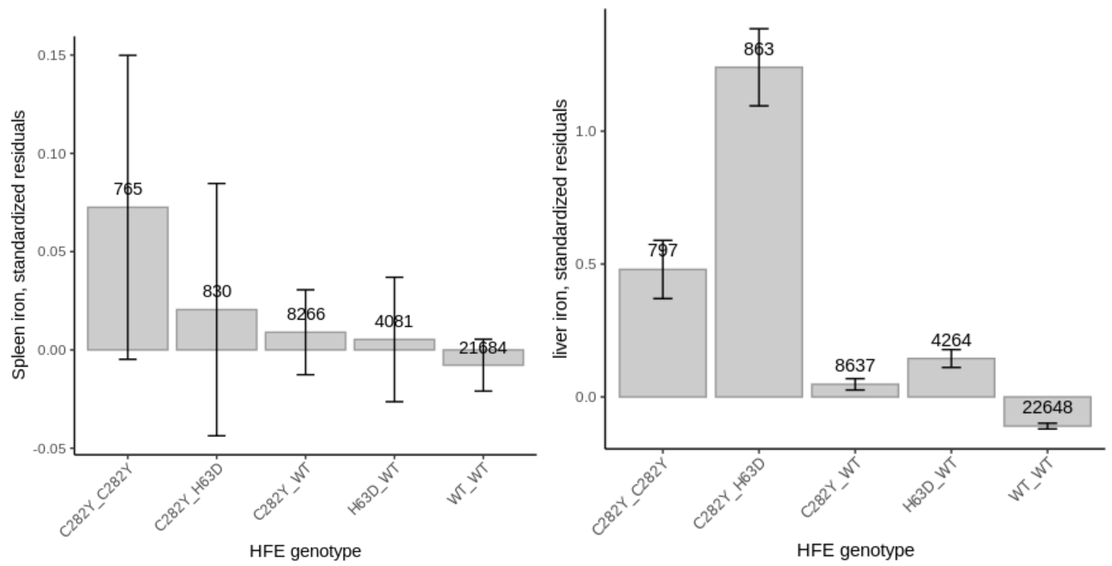


**Supplementary Figure 9: Spleen iron in HFE carriers, homozygotes, compound heterozygotes and non-carriers.** (A) HFEp.Cys282Tyr homozygosity, HFEp.Cys282Tyr/p.His63Asp compound heterozygosity, and HFEp.Cys282Tyr carrier status are not associated with significant differences in spleen iron. Standardized residuals of liver iron content, following adjustment for age, genetic sex, study center, scan date and time (B) Carrier status at the HFE locus affects liver iron. Standardized residuals of spleen iron, following adjustment for age, genetic sex, study center, scan date and time.

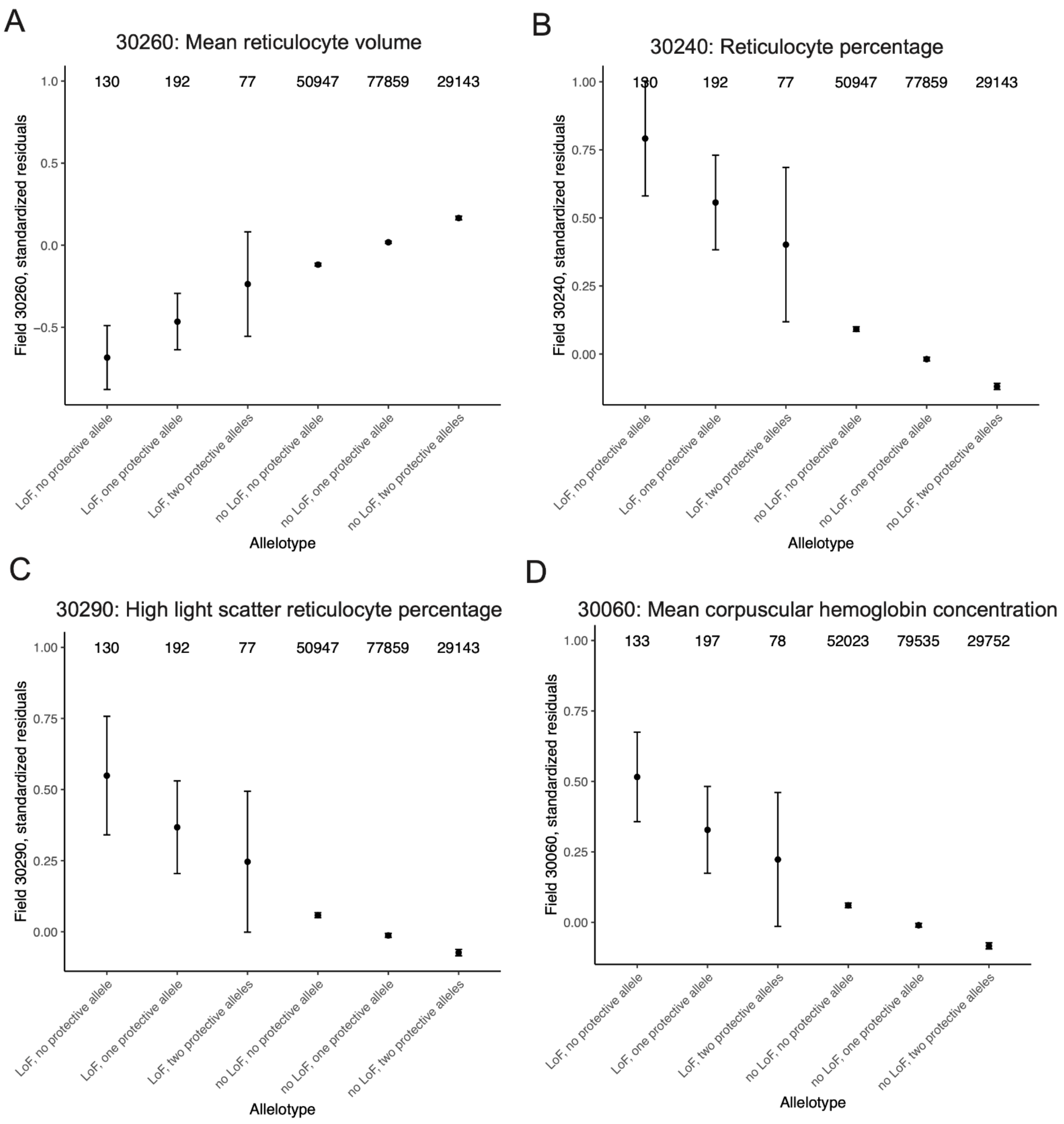


**Supplementary Figure 10**: Allelotype analysis comparing individuals with and without the common, 'protective' alleles in *SPTA1* or *ANK1*, and with and without putative deleterious alleles in one of six hereditary spherocytosis (HS) genes. Standardized residuals, adjusted for age, sex, and global principal components of ancestry are shown for four red blood cell parameters: (A) mean reticulocyte volume (B) reticulocyte percentage (C) high light scatter reticulocyte percentage, (D) mean corpuscular hemoglobin concentration. Sample sizes are shown at top. 95% confidence intervals around group means are shown.

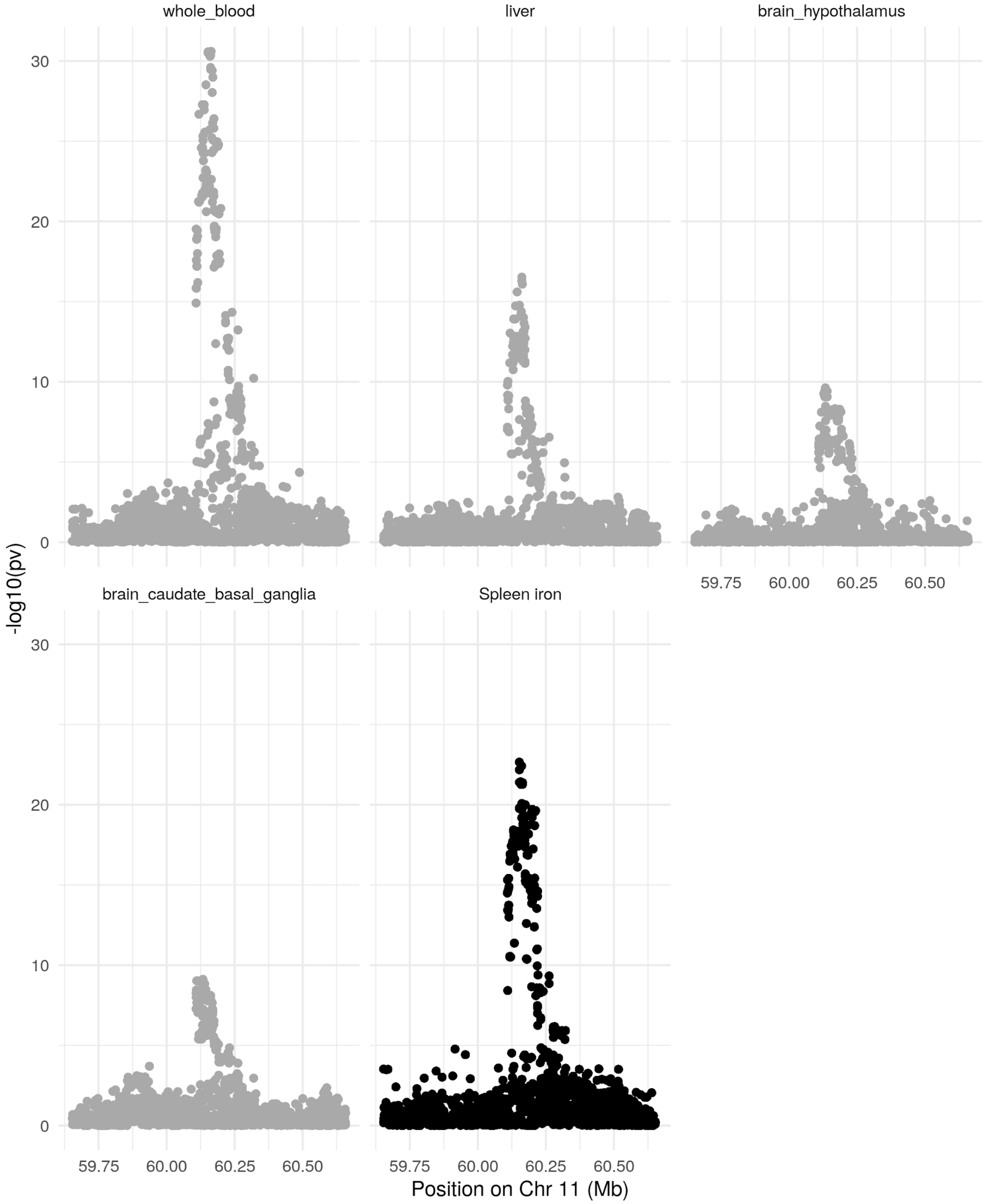


**Supplementary Figure 11: Colocalization of spleen iron and expression of MS4A14 in various tissues**. Tissues are ordered by the strength of the colocalization.
